# Behavioral effects of multiple-dose oxytocin treatment in autism: A randomized, placebo-controlled trial with long-term follow-up

**DOI:** 10.1101/19012112

**Authors:** Sylvie Bernaerts, Bart Boets, Guy Bosmans, Jean Steyaert, Kaat Alaerts

## Abstract

**Background:** Intranasal administration of the ‘prosocial’ neuropeptide oxytocin is increasingly explored as a potential treatment for targeting the core characteristics of autism spectrum disorder (ASD). However, long-term follow-up studies, evaluating the possibility of long-lasting retention effects are currently lacking.

**Methods:** Using a double-blind, randomized, placebo-controlled, parallel design, this pilot clinical trial explored the possibility of long-lasting behavioral effects of four weeks of intranasal oxytocin treatment (24 International Units once daily in the morning) in 40 adult men with ASD. To do so, self-report and informant-based questionnaires assessing core autism symptoms and characterizations of attachment were administered at baseline, immediately after four weeks of treatment (approximately 24 hours after the last nasal spray administration), and at two follow-up sessions, four weeks and one year post-treatment.

**Results:** No treatment-specific effects were identified in the primary outcome assessing social symptoms (Social responsiveness scale, self- and informant-rated). In particular, with respect to self-reported social responsiveness, improvements were evident both in the oxytocin and in the placebo group, yielding no significant between-group difference (p= .37). Also informant-rated improvements in social responsiveness were not significantly larger in the oxytocin, compared to the placebo group (between-group difference: p= .19).

Among the secondary outcome measures, treatment-specific improvements were identified in the Repetitive Behavior Scale and State Adult Attachment Measure, indicating reductions in self-reported repetitive behaviours (p= .04) and reduced feelings of avoidance towards others (p= .03) in the oxytocin group compared to the placebo group, up to one month and even one year post-treatment. Treatment-specific effects were also revealed in screenings of mood states (Profile of Mood States), indicating higher reports of ‘vigor’ (feeling energetic, active, lively) in the oxytocin, compared to the placebo group (p= .03).

**Conclusions:** While no treatment-specific improvements were evident in terms of core social symptoms, the current observations of long-term beneficial effects on repetitive behaviors and feelings of avoidance are promising and suggestive of a therapeutic potential of oxytocin treatment for ASD. However, given the exploratory nature of this pilot study, future studies are warranted to evaluate the long-term effects of OT administration further.

**Trial Registration:** The trial was registered with the European Clinical Trial Registry (Eudract 2014-000586-45) on January 22, 2014 (https://www.clinicaltrialsregister.eu/ctr-search/trial/2014-000586-45/BE).

## 1. Background

Autism Spectrum Disorder (ASD) is characterized by lifelong impairments in social and communicative functioning, and the presence of stereotyped behaviors and interests (1). In the past decade, intranasal administration of the neuropeptide oxytocin (OT) has increasingly been explored as a potential pharmacological treatment for targeting the core characteristics of ASD. Endogenous OT is synthesized in the hypothalamus where neurons of the paraventricular nuclei project to various areas of the central nervous system involved in complex (social) behaviors (e.g. amygdala). In typically developing individuals, OT has been linked to interpersonal bonding, parental care, and the ability to establish trust and form social attachments (2,3).

With respect to ASD, initial clinical trials have demonstrated that a single dose of OT can induce behavioral enhancements on tasks assessing repetitive behavior (4), affective speech comprehension (emotional intonations) (5), facial emotion recognition (6) and social decision making (cyberball computer game) (7). Multiple-dose trials assessing the effects of OT treatment in adults with ASD have also shown beneficial effects. Anagnostou et al. (8) assessed the safety and efficacy of six weeks of intranasal OT treatment on core autism symptom domains (social cognition/function and repetitive behaviors) in 19 adults with ASD (16 men, 3 women), and showed improved emotion recognition, quality of life and tentative improvements in repetitive behaviors after OT treatment. In another study with 20 adult men with ASD, Watanabe et al. (9) studied the effects of six weeks of intranasal OT administration on core autism characteristics and showed significant improvements in social reciprocity and social functioning (social-judgement task). In a more recent large-scale trial, Yamasue et al. (10) also assessed the effects of six weeks of intranasal OT treatment on core autism symptom domains in 106 adult men with ASD and showed significant improvements in repetitive behaviors.

However, a more mixed pattern of results emerged from studies assessing the effects of multiple-dose OT treatment in children with ASD. For example, Dadds et al. (11) assessed behavioral effects of a four-day intranasal OT treatment in 38 boys with ASD (7 to 16 years old) during parent-child interaction training, but found no OT-specific improvements in repetitive behaviors or social responsiveness. Later, also Guastella et al. (12) failed to show beneficial effects after an eight-week OT treatment on core ASD characteristics in slightly older boys (12 to 18 years old). On the other hand, a more recent trial assessing the effects of five weeks of intranasal OT treatment on core ASD characteristics in 31 children with ASD (27 boys, 4 girls) was able to demonstrate improved social responsiveness as reported by the parents (13). Similarly, Parker et al. (14) investigated whether a four-week intranasal OT treatment could improve core autism characteristics in 32 children (6-12 years old) with ASD and showed an increase in parent-reported social responsiveness. Taken together, findings of these initial multiple-dose trials provided indications for a more consistent (positive) pattern of results for OT trials with adults with ASD (8–10), compared to trials with children with ASD (11–14).

To date, however, potential long-term effects of OT treatment that outlast the period of actual administration have not yet been addressed in adults with ASD. These assessments, however, would be of high relevance since repeated administrations over an extended period might induce long-lasting, experience-dependent adaptations within neural circuits. With respect to ASD, it has been postulated that early-life impairments in social attention/orienting may deprive patients of adequate social learning experiences that normally drive the typical development of social brain networks (15). Considering that OT is hypothesized to increase the saliency of social cues (16) OT therapy might induce an enrichment of social experiences that stimulates long-term adaptations in social behaviors.

In this pilot study, we assessed - for the first time - the possibility of long-term retention effects of four weeks of intranasal OT administration on core autism symptom domains (including social responsiveness and restricted and repetitive behaviors), attachment characteristics and general aspects of quality of life in adult men with ASD. While prior multiple-dose trials mainly explored the effects of treatment on these core autism symptom domains, a recent study from our lab (17), showed that two weeks of OT treatment in typically developing young-adult men reduced self-reported feelings of attachment avoidance and increased self-reported feelings of secure attachment toward peers. Against this background and given the growing body of research demonstrating an important role of the oxytocinergic system in interpersonal trust and attachment (18–20), the current study also included an initial assessment of attachment measures to evaluate the long-term effects of OT treatment in adults with ASD. With respect to ASD, Rutgers et al. (21) suggested that most children with ASD (53%) are able to form a secure attachment to caregivers, but that they are significantly less likely to do so compared to typically developing children. In addition, a recent meta-analysis concluded that more severe autism characteristics are associated with less secure attachment in children with ASD (22).

The effects of treatment on core autism characteristics and attachment were assessed immediately after the four-week treatment period, and at follow-up sessions, four weeks and one year post-treatment, to assess the possibility of retention effects that outlast the period of actual administration. In line with prior multiple-dose studies (8,9,17), we hypothesized that multiple doses of OT would improve self-report and informant-based ratings of social responsiveness and repetitive behaviors, and ameliorate self-ratings of attachment characteristics and quality of life. A key question was to evaluate whether any beneficial effects of multiple-dose OT treatment would outlast the period of actual administration until one month (four weeks) and/or one year post-treatment.

## 2. Materials and Methods

### 2.1. General study design

This two-arm, double-blind, randomized, placebo-controlled parallel study was performed at the Leuven University Hospital (Leuven, Belgium) to assess multiple-dose effects of intranasal oxytocin (OT) administration on core autism characteristics and experience of attachment in male adults with ASD. A specific aim of this pilot clinical trial was to assess whether any treatment-induced effects would outlast the period of actual administration. To do so, changes-from-baseline (T0) in self-report and informant-based questionnaire scores were assessed immediately after four weeks of OT treatment (T1), and at two follow-up sessions, four weeks (T2) and one year post-treatment (T3) (See **Figure 1**, CONSORT Flow diagram for number of participants randomized and analysed). Written informed consent was obtained from all participants prior to the study. Consent forms and study design were approved by the local Ethics Committee for Biomedical Research at the University of Leuven, KU Leuven (S56327) in accordance to The Code of Ethics of the World Medical Association (Declaration of Helsinki). The trial was registered with the European Clinical Trial Registry (Eudract 2014-000586-45) and the Belgian Federal Agency for Medicines and Health products. Note that the data presented in the current report are part of a larger clinical trial in which neural measures (i.e. task-based and resting-state magnetic resonance imaging (MRI)) were assessed in addition to the presented questionnaire data. These MRI modalities are however not part of the current report and will be reported elsewhere (manuscripts in preparation).

**Figure 1.**
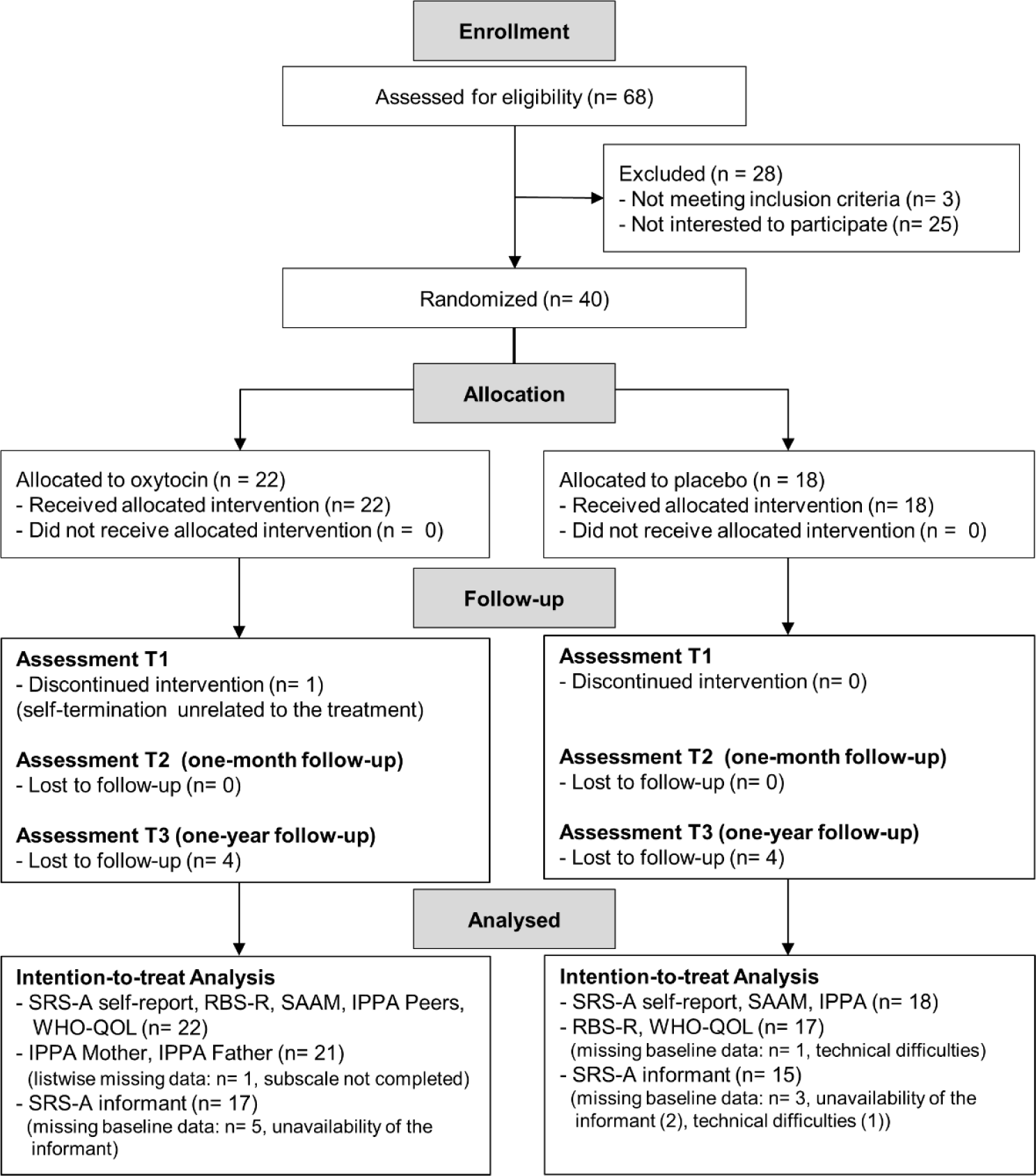
CONSORT Flow diagram. CONSORT Flow diagram. Data were analysed using an intention-to-treat format with last-observations-carried-forward to replace missing data. For participants with missing baseline data, analysis for that measure was excluded list-wise. SRS-A: Social Responsiveness Scale adult version, RBS-R: Repetitive Behavior Scale Revised, SAAM: State Adult Attachment Measure, IPPA: Inventory of Parent and Peer Attachment, WHO-QOL: World Health Organization Quality of Life.

### 2.2. Participants

Forty high-functioning adult men with a formal diagnosis of ASD were recruited between April 2015 and December 2016 from the Autism Expertise Centre at the Leuven University Hospital. The diagnosis was established by a multidisciplinary team (child psychiatrist and/or expert neuropediatrician, psychologist, speech/language pathologist and/or physiotherapist) based on the strict criteria of the DSM-IV-TR (Diagnostic and Statistical Manual of Mental Disorders(1). Prior to the intervention, the Autism Diagnostic Observation Schedule (ADOS or ADOS-2) (23,24) and estimates of intelligence (6-subtest short version of the Wechsler Adult Intelligence Scale-IV Dutch version: Block design, Digit span, Similarities, Vocabulary, Symbol search and Visual puzzles (25) were acquired from all participants (**Table 1**). Inclusion criteria comprised a clinical diagnosis of autism spectrum disorder; gender (male); and age (18-35 years old). Exclusion criteria for participation comprised any neurological disorder (e.g., stroke, epilepsy, concussion); demonstrated genetic disorder; or any contraindication for MRI (note that the MRI data analyses are not part of the current report). Current psychoactive medication use and the presence of comorbid psychiatric disorders were screened (**Supplementary Table S1**). Forty participants were randomly allocated to either the OT (n=22) or the PL group (n=18) (see **Figure 1**, CONSORT Flow diagram). The initial sample size (n=40) was set to be comparable to three prior studies showing significant effects of multiple-dose OT treatment on similar outcome measures (8,9,17).

**Table 1.**
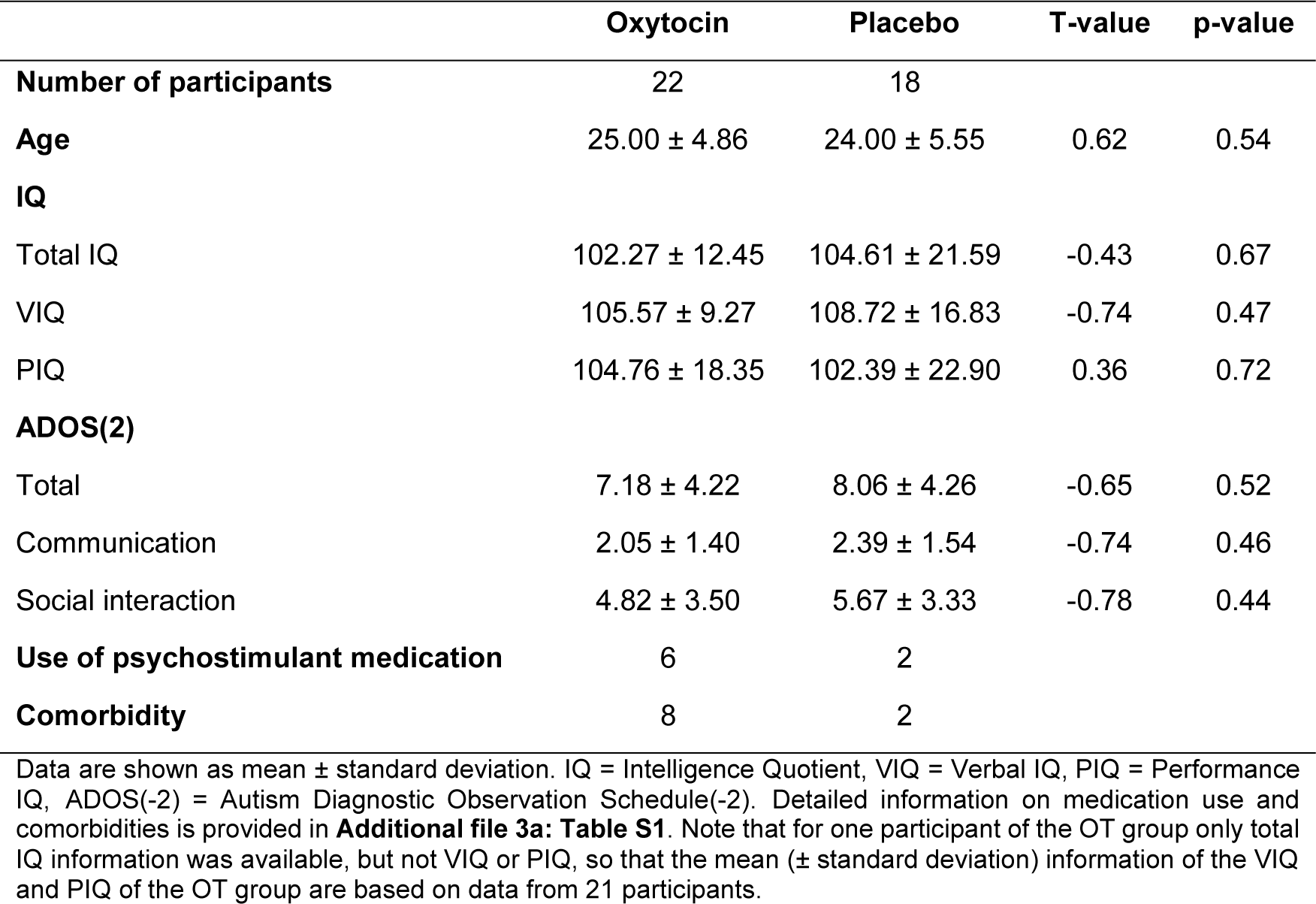
Participant characteristics

|  | Oxytocin | Placebo | T-value | p-value |
| --- | --- | --- | --- | --- |
| **Number of participants** | 22 | 18 |  |  |
| **Age** | 25.00 ± 4.86 | 24.00 ± 5.55 | 0.62 | 0.54 |
| **IQ** |  |  |  |  |
| Total IQ | 102.27 ± 12.45 | 104.61 ± 21.59 | -0.43 | 0.67 |
| VIQ | 105.57 ± 9.27 | 108.72 ± 16.83 | -0.74 | 0.47 |
| PIQ | 104.76 ± 18.35 | 102.39 ± 22.90 | 0.36 | 0.72 |
| **ADOS(2)** |  |  |  |  |
| Total | 7.18 ± 4.22 | 8.06 ± 4.26 | -0.65 | 0.52 |
| Communication | 2.05 ± 1.40 | 2.39 ± 1.54 | -0.74 | 0.46 |
| Social interaction | 4.82 ± 3.50 | 5.67 ± 3.33 | -0.78 | 0.44 |
| **Use of psychostimulant medication** | 6 | 2 |  |  |
| **Comorbidity** | 8 | 2 |  |  |

Data are shown as mean ± standard deviation. IQ = Intelligence Quotient, VIQ = Verbal IQ, PIQ = Performance IQ, ADOS(-2) = Autism Diagnostic Observation Schedule(-2). Detailed information on medication use and comorbidities is provided in **Additional file 3a: Table S1**. Note that for one participant of the OT group only total IQ information was available, but not VIQ or PIQ, so that the mean (± standard deviation) information of the VIQ and PIQ of the OT group are based on data from 21 participants.

### 2.3. Intervention

Participants were assigned to receive the OT or placebo (PL) treatment based on a computer-generated randomized order. Except for the manager of randomization, all research staff conducting the trial, participants and their parents and/or partners were blinded to treatment allocation. OT (Syntocinon®, Sigma-tau) and PL (saline natrium-chloride solution) were administered in amber 15 ml glass bottles with metered pump (ACA Pharma). Each puff per nostril contained 4 international units (IU) of OT. Participants self-administered a daily dose of 24 IU (3 puffs/nostril) over four consecutive weeks (28 doses in total). This dose is in accordance with prior OT administration studies in neurotypical (young) adults (26,27) and adults and adolescents with ASD (6,28,29). At this dosage, no side effects or contraindications of OT have been described (29). All participants received clear instructions about the use of the nasal spray (17,30) and were monitored onsite until approximately two hours after first nasal spray administration. During the course of the treatment, participants were asked to administer the nasal spray in the morning, to keep a daily record of the time point of nasal spray administration, and whether or not they were alone or in company of others the first two hours after administration. Percentage of days at which the spray was administered in the presence of others was not significantly different between treatment groups (OT: 36.8 % (SD 29.9); PL: 36.0 % (SD 25.1); t(37)= .09; p= .93). Participants administered the nasal spray (OT or PL) daily during four consecutive weeks and at the end of each week participants were screened for potential adverse events, side effects (**Supplementary Table S2**) or changes in mood with the Profile of Mood States (POMS) questionnaire (**Table 2** and **Supplementary Figure S1**). Finally, at the end of the trial, participants were asked if they thought they had received OT or PL. The majority of participants thought they had received the PL treatment (79.5%). The proportion of participants that believed they had received the OT treatment was not significantly larger in the actual OT group (28.57%), compared to the PL group (11.11%) (p=.18). Nonetheless, secondary analyses were performed to explore whether treatment effects were modulated by the participants’ own belief about the received treatment (**Supplementary Results**).

**Table 2.**
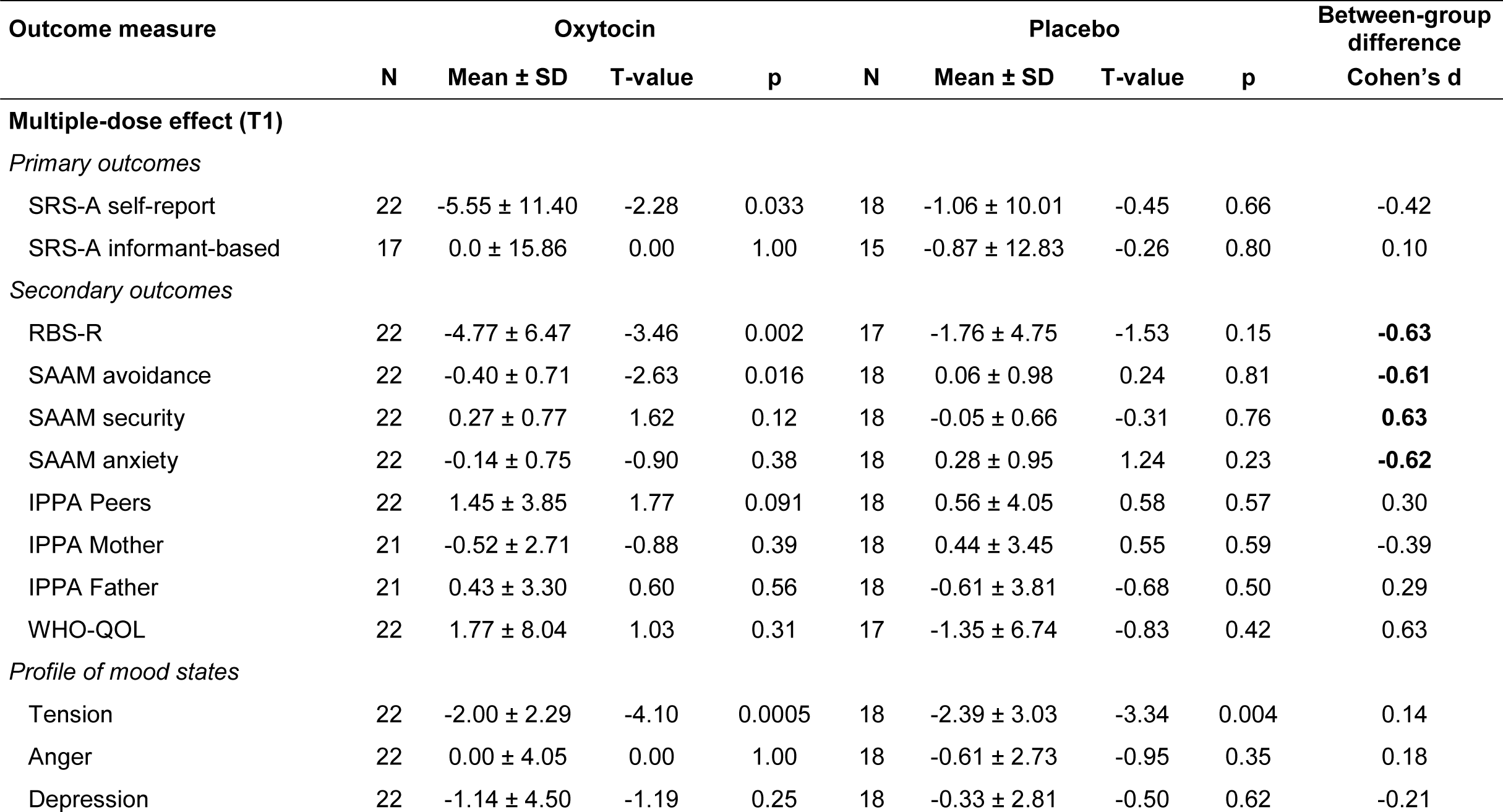
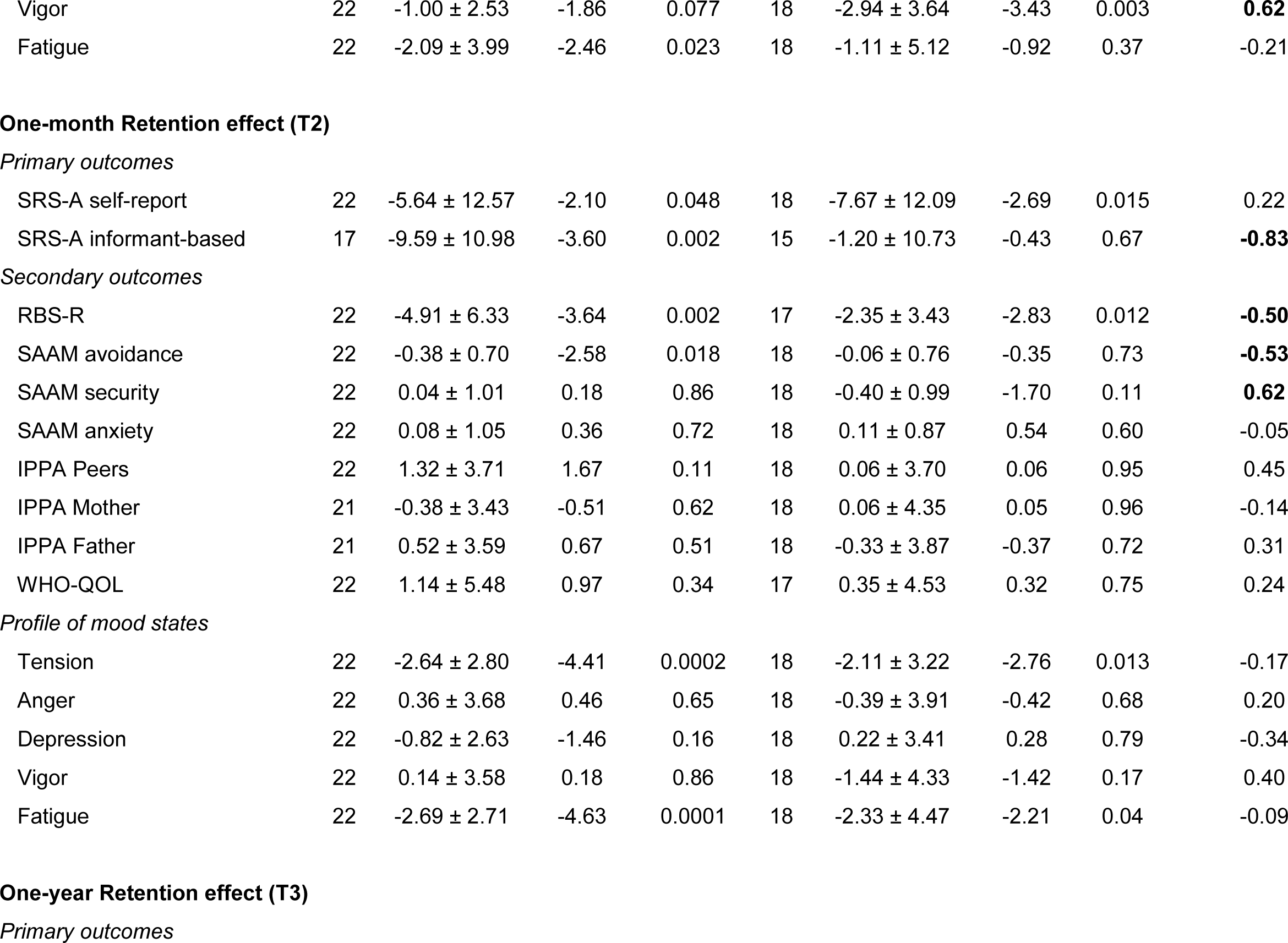
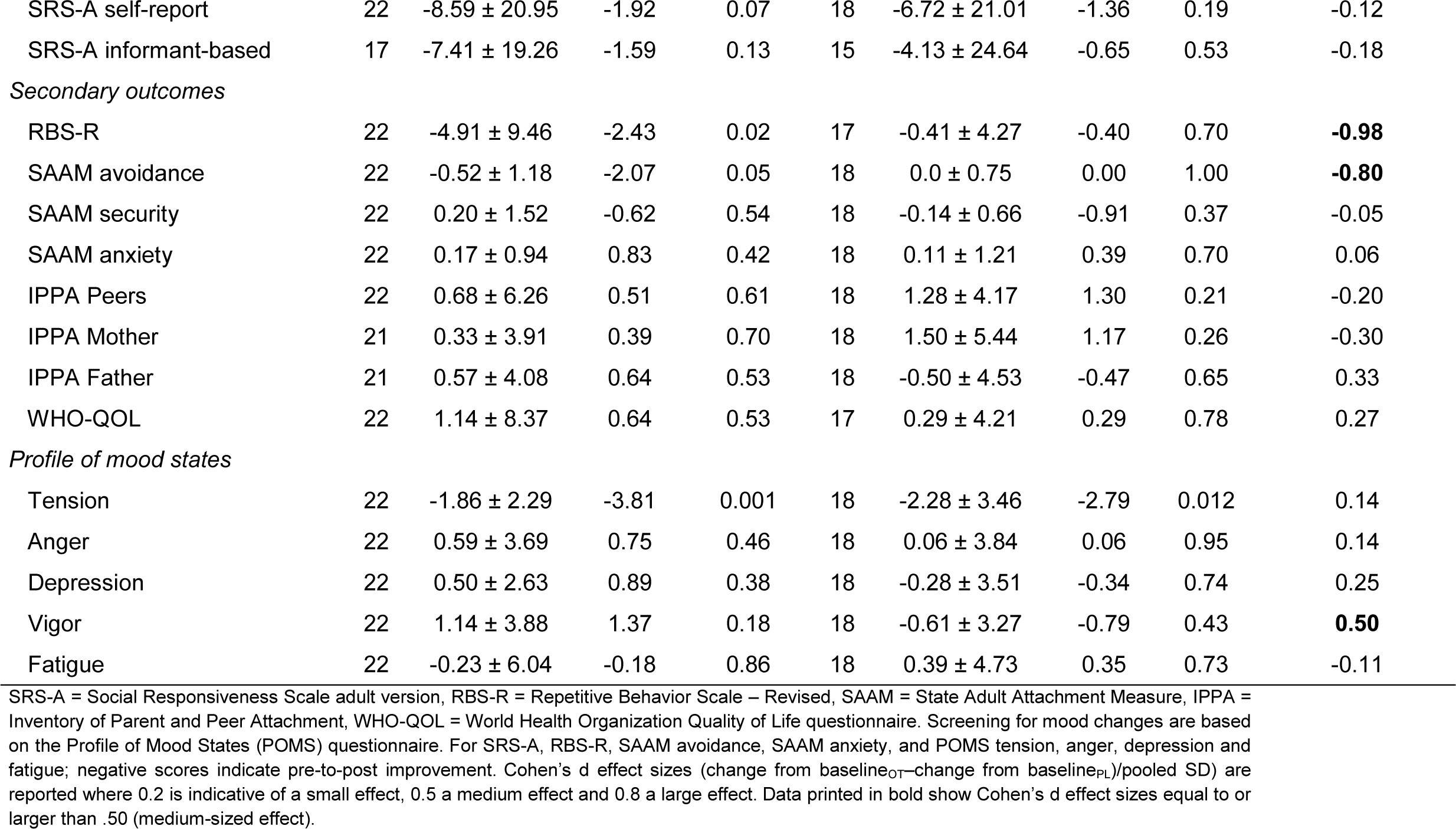
Outcome measures and effects of oxytocin treatment. Mean pre-to-post change scores are listed separately for each treatment group (oxytocin, placebo) and assessment session (T1, T2, T3). Cohen’s d effect sizes of *between-group* differences are reported separately for each outcome measure and assessment session. T- and p-values correspond to single-sample t-tests assessing *within-group* changes from baseline separately for the oxytocin and placebo group.

| Outcome measure | Oxytocin |  |  |  | Placebo |  |  |  | Between-group difference |
| --- | --- | --- | --- | --- | --- | --- | --- | --- | --- |
|  | N | Mean ± SD | T-value | p | N | Mean ± SD | T-value | p | Cohen's d |
| Multiple-dose effect (T1) |  |  |  |  |  |  |  |  |  |
| Primary outcomes |  |  |  |  |  |  |  |  |  |
| SRS-A self-report | 22 | -5.55 ± 11.40 | -2.28 | 0.033 | 18 | -1.06 ± 10.01 | -0.45 | 0.66 | -0.42 |
| SRS-A informant-based | 17 | 0.0 ± 15.86 | 0.00 | 1.00 | 15 | -0.87 ± 12.83 | -0.26 | 0.80 | 0.10 |
| Secondary outcomes |  |  |  |  |  |  |  |  |  |
| RBS-R | 22 | -4.77 ± 6.47 | -3.46 | 0.002 | 17 | -1.76 ± 4.75 | -1.53 | 0.15 | **-0.63** |
| SAAM avoidance | 22 | -0.40 ± 0.71 | -2.63 | 0.016 | 18 | 0.06 ± 0.98 | 0.24 | 0.81 | **-0.61** |
| SAAM security | 22 | 0.27 ± 0.77 | 1.62 | 0.12 | 18 | -0.05 ± 0.66 | -0.31 | 0.76 | **0.63** |
| SAAM anxiety | 22 | -0.14 ± 0.75 | -0.90 | 0.38 | 18 | 0.28 ± 0.95 | 1.24 | 0.23 | **-0.62** |
| IPPA Peers | 22 | 1.45 ± 3.85 | 1.77 | 0.091 | 18 | 0.56 ± 4.05 | 0.58 | 0.57 | 0.30 |
| IPPA Mother | 21 | -0.52 ± 2.71 | -0.88 | 0.39 | 18 | 0.44 ± 3.45 | 0.55 | 0.59 | -0.39 |
| IPPA Father | 21 | 0.43 ± 3.30 | 0.60 | 0.56 | 18 | -0.61 ± 3.81 | -0.68 | 0.50 | 0.29 |
| WHO-QOL | 22 | 1.77 ± 8.04 | 1.03 | 0.31 | 17 | -1.35 ± 6.74 | -0.83 | 0.42 | 0.63 |
| Profile of mood states |  |  |  |  |  |  |  |  |  |
| Tension | 22 | -2.00 ± 2.29 | -4.10 | 0.0005 | 18 | -2.39 ± 3.03 | -3.34 | 0.004 | 0.14 |
| Anger | 22 | 0.00 ± 4.05 | 0.00 | 1.00 | 18 | -0.61 ± 2.73 | -0.95 | 0.35 | 0.18 |
| Depression | 22 | -1.14 ± 4.50 | -1.19 | 0.25 | 18 | -0.33 ± 2.81 | -0.50 | 0.62 | -0.21 |
| Vigor | 22 | -1.00 ± 2.53 | -1.86 | 0.077 | 18 | -2.94 ± 3.64 | -3.43 | 0.003 | **0.62** |
| Fatigue | 22 | -2.09 ± 3.99 | -2.46 | 0.023 | 18 | -1.11 ± 5.12 | -0.92 | 0.37 | -0.21 |
| Primary outcomes |  |  |  |  |  |  |  |  |  |
| SRS-A self-report | 22 | -5.64 ± 12.57 | -2.10 | 0.048 | 18 | -7.67 ± 12.09 | -2.69 | 0.015 | 0.22 |
| SRS-A informant-based | 17 | -9.59 ± 10.98 | -3.60 | 0.002 | 15 | -1.20 ± 10.73 | -0.43 | 0.67 | **-0.83** |
| Secondary outcomes |  |  |  |  |  |  |  |  |  |
| RBS-R | 22 | -4.91 ± 6.33 | -3.64 | 0.002 | 17 | -2.35 ± 3.43 | -2.83 | 0.012 | **-0.50** |
| SAAM avoidance | 22 | -0.38 ± 0.70 | -2.58 | 0.018 | 18 | -0.06 ± 0.76 | -0.35 | 0.73 | **-0.53** |
| SAAM security | 22 | 0.04 ± 1.01 | 0.18 | 0.86 | 18 | -0.40 ± 0.99 | -1.70 | 0.11 | **0.62** |
| SAAM anxiety | 22 | 0.08 ± 1.05 | 0.36 | 0.72 | 18 | 0.11 ± 0.87 | 0.54 | 0.60 | -0.05 |
| IPPA Peers | 22 | 1.32 ± 3.71 | 1.67 | 0.11 | 18 | 0.06 ± 3.70 | 0.06 | 0.95 | 0.45 |
| IPPA Mother | 21 | -0.38 ± 3.43 | -0.51 | 0.62 | 18 | 0.06 ± 4.35 | 0.05 | 0.96 | -0.14 |
| IPPA Father | 21 | 0.52 ± 3.59 | 0.67 | 0.51 | 18 | -0.33 ± 3.87 | -0.37 | 0.72 | 0.31 |
| WHO-QOL | 22 | 1.14 ± 5.48 | 0.97 | 0.34 | 17 | 0.35 ± 4.53 | 0.32 | 0.75 | 0.24 |
| Profile of mood states |  |  |  |  |  |  |  |  |  |
| Tension | 22 | -2.64 ± 2.80 | -4.41 | 0.0002 | 18 | -2.11 ± 3.22 | -2.76 | 0.013 | -0.17 |
| Anger | 22 | 0.36 ± 3.68 | 0.46 | 0.65 | 18 | -0.39 ± 3.91 | -0.42 | 0.68 | 0.20 |
| Depression | 22 | -0.82 ± 2.63 | -1.46 | 0.16 | 18 | 0.22 ± 3.41 | 0.28 | 0.79 | -0.34 |
| Vigor | 22 | 0.14 ± 3.58 | 0.18 | 0.86 | 18 | -1.44 ± 4.33 | -1.42 | 0.17 | 0.40 |
| Fatigue | 22 | -2.69 ± 2.71 | -4.63 | 0.0001 | 18 | -2.33 ± 4.47 | -2.21 | 0.04 | -0.09 |
| Primary outcomes |  |  |  |  |  |  |  |  |  |
| SRS-A self-report | 22 | -8.59 ± 20.95 | -1.92 | 0.07 | 18 | -6.72 ± 21.01 | -1.36 | 0.19 | -0.12 |
| SRS-A informant-based | 17 | -7.41 ± 19.26 | -1.59 | 0.13 | 15 | -4.13 ± 24.64 | -0.65 | 0.53 | -0.18 |
| *Secondary outcomes* |  |  |  |  |  |  |  |  |  |
| RBS-R | 22 | -4.91 ± 9.46 | -2.43 | 0.02 | 17 | -0.41 ± 4.27 | -0.40 | 0.70 | **-0.98** |
| SAAM avoidance | 22 | -0.52 ± 1.18 | -2.07 | 0.05 | 18 | 0.0 ± 0.75 | 0.00 | 1.00 | **-0.80** |
| SAAM security | 22 | 0.20 ± 1.52 | -0.62 | 0.54 | 18 | -0.14 ± 0.66 | -0.91 | 0.37 | -0.05 |
| SAAM anxiety | 22 | 0.17 ± 0.94 | 0.83 | 0.42 | 18 | 0.11 ± 1.21 | 0.39 | 0.70 | 0.06 |
| IPPA Peers | 22 | 0.68 ± 6.26 | 0.51 | 0.61 | 18 | 1.28 ± 4.17 | 1.30 | 0.21 | -0.20 |
| IPPA Mother | 21 | 0.33 ± 3.91 | 0.39 | 0.70 | 18 | 1.50 ± 5.44 | 1.17 | 0.26 | -0.30 |
| IPPA Father | 21 | 0.57 ± 4.08 | 0.64 | 0.53 | 18 | -0.50 ± 4.53 | -0.47 | 0.65 | 0.33 |
| WHO-QOL | 22 | 1.14 ± 8.37 | 0.64 | 0.53 | 17 | 0.29 ± 4.21 | 0.29 | 0.78 | 0.27 |
| *Profile of mood states* |  |  |  |  |  |  |  |  |  |
| Tension | 22 | -1.86 ± 2.29 | -3.81 | 0.001 | 18 | -2.28 ± 3.46 | -2.79 | 0.012 | 0.14 |
| Anger | 22 | 0.59 ± 3.69 | 0.75 | 0.46 | 18 | 0.06 ± 3.84 | 0.06 | 0.95 | 0.14 |
| Depression | 22 | 0.50 ± 2.63 | 0.89 | 0.38 | 18 | -0.28 ± 3.51 | -0.34 | 0.74 | 0.25 |
| Vigor | 22 | 1.14 ± 3.88 | 1.37 | 0.18 | 18 | -0.61 ± 3.27 | -0.79 | 0.43 | **0.50** |
| Fatigue | 22 | -0.23 ± 6.04 | -0.18 | 0.86 | 18 | 0.39 ± 4.73 | 0.35 | 0.73 | -0.11 |

SRS-A = Social Responsiveness Scale adult version, RBS-R = Repetitive Behavior Scale – Revised, SAAM = State Adult Attachment Measure, IPPA = Inventory of Parent and Peer Attachment, WHO-QOL = World Health Organization Quality of Life questionnaire. Screening for mood changes are based on the Profile of Mood States (POMS) questionnaire. For SRS-A, RBS-R, SAAM avoidance, SAAM anxiety, and POMS tension, anger, depression and fatigue; negative scores indicate pre-to-post improvement. Cohen's d effect sizes (change from baseline<sub>OT</sub>—change from baseline<sub>PL</sub>)/pooled SD) are reported where 0.2 is indicative of a small effect, 0.5 a medium effect and 0.8 a large effect. Data printed in bold show Cohen's d effect sizes equal to or larger than .50 (medium-sized effect).

### 2.4. Outcome Measures

The Social Responsiveness Scale (for adults) (SRS-A) total score was used as the primary outcome measure (self-report and informant-based versions). The other behavioral questionnaires were considered secondary: Repetitive Behavior Scale - Revised (RBS-R); State Adult Attachment Measure (SAAM); Inventory of Parent and Peer Attachment (IPPA); Quality of Life questionnaire of the World Health Organization (WHO-QOL) (all self-report). More detailed information on each of the adopted questionnaires is provided in **Supplementary Methods**.

In short, the SRS-A (64 items) (31) comprises four subscales examining social communication, social awareness, social motivation and rigidity/repetitiveness, using a four-point Likert-scale. SRS-A raw total scores were adopted.

The RBS-R (43 items) (32) examines a heterogeneous set of repetitive behaviors including stereotypic behavior, self-injurious behavior, compulsive behavior, ritualistic behavior, sameness behavior and restricted interests behavior, using a four-point Likert-scale. RBS-R raw total scores were adopted.

The SAAM (33) comprises three subscales examining attachment security (e.g., “I feel like I have someone to rely on”) (7 items); attachment anxiety (e.g., “I feel a strong need to be unconditionally loved right now”) (7 items); and attachment avoidance (e.g., “If someone tried to get close to me, I would try to keep my distance”) (7 items) using a seven-point Likert-scale. SAAM raw subscale scores were adopted.

The IPPA (34) examines trait attachment to (i) mother (12 items); (ii) father (12 items); (iii) peers (12 items), using a four-point Likert-scale. The IPPA assesses attachment security along three dimensions: degree of mutual trust, quality of communication and extent of anger and alienation. IPPA raw subscale scores were adopted.

Finally, the abbreviated version of the WHO-QOL (35) assesses general quality of life related to physical health, psychological health, social relationships, and environment using a five-point Likert scale. WHO-QOL raw total scores were adopted.

All questionnaires were assessed at baseline (T0), immediately after four consecutive weeks of nasal spray administration (approximately 24 hours after the last nasal spray administration) (T1), and at follow-up sessions, four weeks (T2) and one year post-treatment (T3). Given that this is an exploratory pilot study evaluating the possibility of long-term retention effects of OT treatment, no primary time point was pre-specified.

### 2.5. Data Analysis

For each questionnaire, baseline differences between groups were assessed using two-sample t-tests. To assess *between-group differences*, pre-to-post difference scores were calculated for each assessment session (T1, T2, T3) and difference scores were subjected to a linear mixed-effects model (one-tailed) with the random factor ‘subject’, and the fixed factors ‘treatment’ (OT, PL), ‘session’ (T1, T2, T3) and ‘treatment x session’ interaction. An intention-to-treat format and last-observations-carried-forward to replace missing data was adopted. For participants with missing baseline data, analysis for that measure was excluded list-wise (**Figure 1** CONSORT Flow diagram). Cohen’s d effect sizes of between-group differences (change from baselineOT–change from baselinePL)/pooled standard deviation) are reported in **Table 2**, separately for each outcome measure and assessment session, where 0.2 is indicative of a small effect, 0.5 a medium effect and 0.8 a large effect (36). Additionally, pre-to-post difference scores were subjected to single-sample t-tests to assess within-group changes (compared to baseline) in the OT group and PL group separately (**Table 2**). All statistics were executed with Statistica 8 (StatSoft. Inc. Tulsa, USA). Given that this is an exploratory pilot study evaluating long-term effects of OT treatment, the significance level was set at p < .05 for all analyses, without correction for multiple comparisons.

## 3. Results

No significant baseline differences were revealed between participants allocated to the OT or PL group for any of the questionnaires (see **Supplementary Table S3**) or in terms of participant characteristics (see **Table 1**).

### Primary outcome - Social Responsiveness Scale (SRS-A)

#### Self-rated SRS-A

Between-group analyses revealed no significant main effect of treatment (F(1,76)= .12, p= .37, ŋ^2^ = .00), nor a treatment x session interaction effect (F(2,76)= 1.17, p= .16, ŋ^2^ = .03), indicating that pre-to-post changes in self-reported social responsiveness were not significantly larger in the OT compared to the PL group (see **Figure 2** and **Table 2** for the effect sizes of between-group differences separately for each session). Within-group analyses showed that SRS-A scores were significantly reduced (compared to baseline) in the OT group at session T1 (immediately after treatment: p= .033), T2 (one month post-treatment; p= .048) and at trend-level at session T3 (one year post-treatment: p= .07). However, a similar reduction (compared to baseline) was evident in the PL group at session T2 (p= .015), indicating no specific benefit of OT over the PL treatment (see **Table 2** reporting single-sample t-tests assessing within-group changes from baseline).

**Figure 2.**
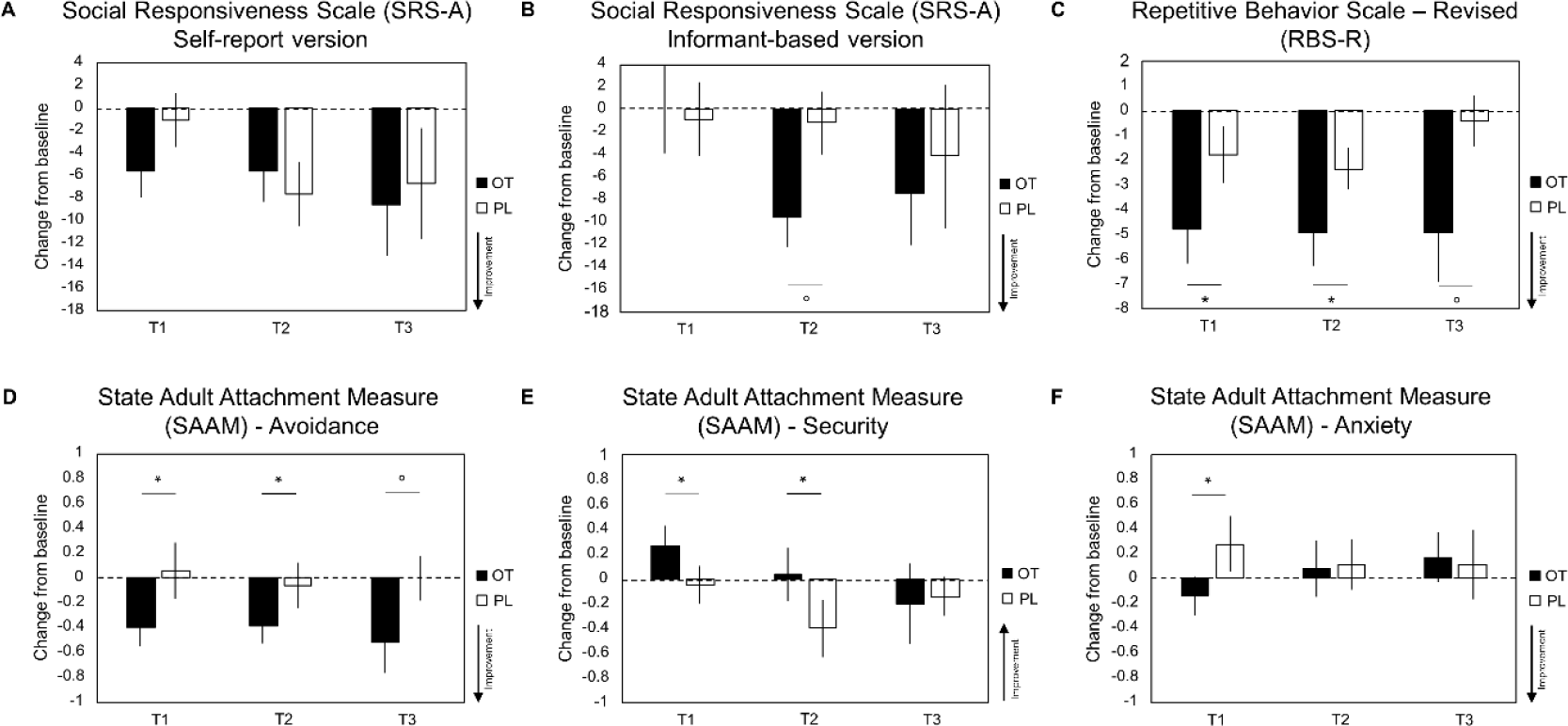
Effects of oxytocin treatment on autism symptoms and attachment. Mean pre-to-post changes (change from baseline) on self-report and informant-based questionnaires are visualized for the oxytocin (OT) and placebo (PL) treatment groups at assessment session ‘T1’ (immediately after the four-week treatment), ‘T2’ (at follow-up, one month post-treatment) and ‘T3’ (at follow-up, one year post-treatment). Mean changes from baseline are visualized separately for (A) Social Responsiveness Scale (SRS-A) self-report version, (B) SRS-A informant-based version, (C) Repetitive Behavior Scale – Revised (RBS-R), (D) State Adult Attachment Measure (SAAM) Avoidance subscale, (E) SAAM Security subscale, and (F) SAAM Anxiety subscale. Lower scores indicate improvement for the SRS-A, RBS-R, SAAM Avoidance and SAAM Anxiety questionnaires. For the SAAM Security questionnaire, higher scores indicate improvement. Vertical bars denote +/- standard errors. Asterisks (*) indicate Cohen’s d ≥ .50 (medium-sized effect). Circles (°) indicate Cohen’s d ≥.80 (large-sized effect).

#### Informant-rated SRS-A

Between-group analyses of the informant-rated SRS-A scores revealed no significant main effect of treatment (F(1, 60)= .78, p= .19, ŋ^2^ = .03), nor a treatment x session interaction effect (F(2, 60)= .83, p= .22, ŋ^2^ = .03), indicating that pre-to-post changes in informant-rated social responsiveness were not significantly larger in the OT compared to the PL group (see **Figure 2** and **Table 2** for the effect sizes of between-group differences separately for each session). Note however that at session T2 (one month post-treatment), informant-rated SRS-A scores were significantly reduced in the OT group (compared to baseline) (p= .002), but not in the PL group (p= .67) (see **Table 2** reporting single-sample t-tests assessing within-group changes from baseline).

### Secondary outcome - Repetitive Behavior Scale – Revised (RBS-R)

In terms of repetitive behaviors, between-group analyses identified a significant main effect of treatment (F(1, 74)= 3.20, p= .04, ŋ^2^ = .08) (but no treatment x session interaction: F(2, 74)= 1.04, p= .18, ŋ^2^ = .03), indicating that across assessment sessions, pre-to-post improvements in repetitive behaviors were significantly larger in the OT compared to the PL group (see **Figure 2** and **Table 2** for the effect sizes of between-group differences separately for each session). Within-group analyses confirmed that RBS-R scores were significantly reduced (compared to baseline) in the OT group at session T1 (immediately after treatment: p= .002), T2 (one month post-treatment; p= .002) and session T3 (one year post-treatment: p= .02), but not consistently in the PL group (T1: p= .15; T2: p= .012; T3: p=.70) (see **Table 2** reporting single-sample t-tests assessing within-group changes from baseline).

### Secondary outcome - State adult attachment measure (SAAM)

#### Attachment Avoidance

In terms of attachment avoidance, between-group analyses identified a significant main effect of treatment (F(1,76)= 3.70, p= .03, ŋ^2^ = .09) (but no treatment x session interaction: F(2,76)= .27, p= .38, ŋ^2^ = .01), indicating that across assessment sessions, pre-to-post improvements in attachment avoidance were significantly larger in the OT compared to the PL group (see **Figure 2** and **Table 2** and for the effect sizes of between-group differences separately for each session). Within-group analyses confirmed that attachment avoidance scores were significantly reduced (compared to baseline) in the OT group at session T1 (immediately after treatment: p= .016), T2 (one-month post-treatment; p= .018) and session T3 (one-year post-treatment: p= .05), but not in the PL group (T1: p= .81; T2: p= .73; T3: p=1.00) (see **Table 2** reporting single-sample t-tests assessing within-group changes from baseline).

#### Attachment Security

Between-group analyses identified no main effect of treatment F(1,76)= .88, p= .18, ŋ^2^ = .02), nor a treatment x session interaction effect (F(2,76)= 1.08, p= .17, ŋ^2^ = .03), indicating no treatment-specific improvement in attachment security across assessment sessions (see **Figure 2** and **Table 2** for the effect sizes of between-group differences separately for each session). Also no significant within-group pre-to-post changes were identified in the OT or PL group separately (see **Table 2**).

#### Attachment Anxiety

Between-group analyses identified no main effect of treatment (F(1,76)= .25, p= .31, ŋ^2^ = .01), nor a treatment x session interaction effect (F(2,76)= 1.566, p= .10, ŋ^2^ = .04), indicating no treatment-specific improvement in attachment anxiety across assessment sessions (see **Figure 2** and **Table 2** and for the effect sizes of between-group differences separately for each session). Also no significant within-group pre-to-post changes were identified in the OT or PL group separately (see **Table 2**).

### Secondary outcome - Inventory of Parent and Peer Attachment (IPPA)

Between-group analyses showed that pre-to-post changes in self-reported secure attachment towards peers and parents were not significantly larger in the OT compared to the PL group (no main effects of treatment: Peers: F(1,76)= .20, p= .33, ŋ^2^ = .01; Mother: F(1,74)= .57, p= .23, ŋ^2^ = .02; Father: F(1,74)= .78, p= .19, ŋ^2^ = .02; nor treatment x session interaction effects: Peers: F(2,76)= 1.08, p= .17, ŋ^2^ = .03; Mother: F(2,74)= .32, p= .36, ŋ^2^ = .01; Father: F(2,74)= .03, p= .49, ŋ^2^ = .00) (see **Supplementary Figure S2** and **Table 2** for the effect sizes of between-group differences separately for each session). Also no significant within-group pre-to-post changes were identified in the OT or PL group separately (see **Table 2**).

### Secondary outcome - World Health Organization Quality of Life (WHO-QOL) – Bref

Between-group analyses showed that pre-to-post changes in self-reported quality of life were not significantly larger in the OT, compared to the PL group (no main effect of treatment: F(1,74)= .77, p= .19, ŋ^2^ = .02, nor a treatment x session interaction effect: F(2,74)= .96, p= .19, ŋ^2^ = .03) (see **Supplementary Figure S2** and **Table 2** for the effect sizes of between-group differences separately for each session). Also no significant within-group pre-to-post changes were identified in the OT or PL group separately (see **Table 2**).

### Screening of changes in mood and side effects

As listed in detail in **Supplementary Table S2**, only minimal, non-treatment specific side effects were reported.

In terms of changes in mood as assessed with the Profile of Mood states (POMS), between-group analyses identified a significant main effect of treatment for the mood state ‘vigor’ (F(1,76)= 4.09, p= .03, ŋ^2^ = .10) (but no treatment x session interaction: F(2,76)= .04, p= .96, ŋ^2^ = .001), indicating that across assessment sessions, self-reports of ‘vigor’ (feeling ‘energetic’, ‘active’, ‘lively’) were significantly higher in the OT group compared to the PL group (see **Table 2** for the effect sizes of between-group differences separately for each session). While no significant pre-to-post changes were evident within the OT group, the PL group showed a significant reduction (compared to baseline) in self-reported vigor at session T1 (p= .002) (see **Table 2** reporting single-sample t-tests assessing within-group changes from baseline).

No treatment-specific changes were identified for the other mood states (tension, anger, depression, fatigue) (see **Table 2** and **Supplementary Figure S1**), although note that significant reductions (compared to baseline) in feelings of tension and fatigue were reported both in the OT group and in the PL group (see **Table 2** reporting single-sample t-tests assessing within-group changes from baseline).

### Associations between ASD characteristics and attachment characteristics

Taken together, treatment-specific effects of a four-week OT treatment were most pronounced in terms of improvements in repetitive behaviors (RBS-R) and perceived attachment avoidance (SAAM). Here, we specifically explored whether and how the quantitative autism characteristics (SRS-A and RBS-R) (assessed at baseline) were associated with the adopted attachment characteristics (SAAM and IPPA). We additionally explored whether individuals with ASD displayed more impairments in attachment, when their baseline behavioral characterizations were compared to those previously obtained from a sample of neurotypical individuals (n=40, mean age = 21.1, S.D. = 2.6) (data adopted from (17)).

Higher self-reported SRS-A scores (at baseline) (more impairment in social responsiveness) were significantly associated with lower perceived secure attachment (IPPA) towards peers (r= -.55, p< .001), mother (r= -.51, p= .001) and father (r= -.34, p= .034) and with higher perceived attachment avoidance (SAAM) (r= .38, p= .018), but not with other reports of attachment characteristics (security: r= -.26, p= .12; anxiety: r= .08, p= .63). Further, higher scores on the RBS-R (more frequent and/or severe repetitive and restricted behaviors) were significantly associated with lower perceived secure attachment (IPPA) towards the mother (r=-.56, p< .001), but not the father (r= -.31; p= .056) or peers (r= -.22, p= .19). Finally, higher scores on the RBS-R were also significantly associated with higher perceived attachment avoidance (r= .50, p= .002), but not with other reports of attachment characteristics (security: r= -.11, p= .50; anxiety: r= -.02, p= .89).

Notably, exploratory analyses also showed that as a group, the individuals with ASD reported significantly higher perceived attachment avoidance (t(76)=-2.51, p=.014), lower attachment security (t(76)=2.48, p=.015), and a trend towards lower perceived secure attachment towards peers (IPPA) (t(76)=1.74, p=.085), when compared to a neurotypical sample of adult men (data obtained from (17)) (**Supplementary Table S4**).

## 4. Discussion

The current trial demonstrated no treatment-specific effects in the primary outcome assessing social symptoms (SRS-A, self- and informant-rated). In particular, with respect to self-reports of social responsiveness, pre-to-post improvements were evident both in the OT (at session T1 and T2) and in the PL group (at session T2), but with no specific benefit of OT over the PL treatment (no significant between-group difference). Correspondingly, in three previous long-term administration trials, improvements in social symptoms (SRS-A: 9, 12) (ADOS subscale assessing social reciprocity: 10) were identified both in participants receiving the OT treatment and in those receiving the placebo treatment, but with no significant difference between the two groups. As suggested by Yatawara et al. (2016), one explanation for these unspecific effects may be the presence of a placebo response which has been shown to occur frequently in paediatric autism pharmacological and dietary placebo-controlled trials (40). In the context of OT trials, increased public attention over the last decade may have influenced the expectations of patients or parents especially with respect to the anticipated effects of OT treatment on social functioning (hence the observed unspecific improvements in self-rated SRS-A scores directly assessing the social domain).

Also with respect to the effect of multiple-dose OT treatment on informant-based reports of social responsiveness (SRS-A), no overall treatment-specific improvements were observed (despite the identification of a reduction in informant-rated symptom severity in the OT group, but not in the PL group at session T2) (see **Table 2** reporting within-group changes). In prior studies with young children with ASD, significant treatment-specific improvements in caregiver-rated social responsiveness have been identified immediately after 4 or 5 weeks of OT treatment (13,14). The lack of a treatment-specific effect in our trial versus previous trials in children might relate to the frequency of contact of children versus adult populations with their respective informants (i.e., more frequent, sustained child-informant contact), potentially rendering more subtle improvements in social functioning in the adult population to remain undetected by informants.

While no significant treatment-specific effects were identified in social symptoms, exploratory analyses identified long-lasting OT-specific improvements in a secondary outcome assessing attachment characteristics (SAAM), indicating a reduction in feelings of avoidance in the OT, compared to the PL group. While the exact neuromodulatory mechanisms of OT treatment are unknown, OT has been implicated in enhancing the salience of socially-relevant cues, inducing reductions in (social) stress and anxiety, and modulating approach/avoidance motivational tendencies, presumably by impacting on limbic circuits (e.g. amygdala) and the central reward system (e.g. nucleus accumbens). As such, by enhancing social salience and reducing social stress/anxiety, the daily OT administrations over a course of four weeks, may have induced increased feelings of approachability (reduced avoidance) during social interactions. Furthermore, the observation that these beneficial effects of multiple-dose OT treatment on perceived attachment avoidance outlasted the period of actual administration until one month and one year post-treatment, provides support to the notion that repeated administrations over an extended period of time might induce long-lasting adaptations in social brain circuits, presumably in an experience-dependent manner. Indeed, through positive re-enforcement, the recursive experience of the social environment as more ‘secure’ or ‘approachable’ (during the period of actual OT administrations) can be anticipated to have contributed to the observed long-lasting adaptations in one’s motivational tendencies (i.e. increased feelings of social approachability). Considering that attachment avoidance reflects a reluctance to trust others and an emphasis on autonomy, whereas attachment anxiety reflects insecurity about oneself (low trust in oneself) and fear of being rejected (33), our results suggest that the four-week OT treatment predominantly improved a person’s reluctance towards closeness or trust in others (i.e., attachment avoidance), but that it could not induce significant long-term alterations in a person’s feelings of insecurity about one’s own abilities (i.e., attachment anxiety). The notion that OT may thus predominantly influence one’s reluctance to engage in closeness or intimacy with others (rather than one’s fear of being rejected) may be interpreted within the framework of the recently proposed affiliative-motivation hypothesis (37) suggesting that OT specifically acts by increasing affiliative strivings and that individuals with a decreased tendency to affiliate (e.g. avoidantly attached individuals rather than anxiously attached individuals) may be most likely to benefit from OT treatment. Overall, the observation of a beneficial effect of OT on feelings of approachability is in line with findings from a previous study from our lab (17), in which we demonstrated similar treatment-specific reductions in perceived avoidant attachment (as well as improvements in perceived secure attachment to peers) after a two-week course of OT treatment in neurotypical men. Our findings also extend previous studies showing beneficial effects of a single dose of OT on attachment security (20), development of trust and cooperation (18) and improved communal traits and altered agency (19).

In addition to the treatment-specific effect on attachment avoidance, long-lasting treatment-specific improvements were also identified in terms of repetitive behaviors (RBS-R), indicating an overall reduction in repetitive behaviors in the OT group, compared to the PL group. While the exact link between expressions of repetitive behaviors and difficulties in the social domain is unclear, it has been suggested that at least in a subset of individuals with ASD, the experience of the external (social) milieu as ‘unapproachable’ or even ‘threatening’ may result in an increased ‘need for sameness’ in order to sustain a level of control over the external surroundings (38,39). The current study provides preliminary evidence that multiple-dose OT treatment may relieve an individual from this increased ‘need for sameness’ and the resulting need for engaging in repetitive and restricted behaviors. Overall, these observed effects on repetitive behaviors are in line with previous trials with adult men with ASD showing beneficial effects of OT on repetitive behaviors after 4 hours of intravenous OT administration (4) and after 6 weeks of daily administrations (8,10). Note however that one six-week trial with adult men with ASD did not show OT-specific improvements on informant-based reports of repetitive behaviors (9). Also, in previous trials with children and adolescents with ASD, no improvements on repetitive behaviors were observed after 4 days or 4, 5, or 8 weeks of daily OT administration (11–14). In this view, it appears that beneficial effects of OT treatment on repetitive behaviors were mostly demonstrated in studies that adopted assessments based on self-reports (current study, (4,8)), whereas no beneficial effects were evident in studies adopting informant-based reports of repetitive behavior (9–14, with the exception of 10). Together, these findings may therefore indicate that self-reports, as opposed to informant-based reports, may be more sensitive for capturing subtle, self-experienced changes in repetitive behaviors.

While not included as an explicit outcome measure, screenings of changes in mood states (Profile of Mood States questionnaire (POMS)) showed a treatment-specific effect for the mood state ‘vigor’, indicating higher reports of feelings of vigor (e.g., feeling ‘energetic’, ‘active’, ‘lively’) in the OT group, compared to the PL group. To our knowledge, this is the first study adopting screenings of mood states in an OT trial with ASD patients. In a previous study from our lab (17), the POMS was also adopted to evaluate the effects of a two-week OT treatment in neurotypical men, and while here, no treatment-specific changes in vigor were detected, the POMS revealed OT-specific reductions in feelings of tension and anger. In the current study, reductions in feelings of tension were also reported, but irrespective of received treatment, indicating no specific benefit of OT over the PL treatment. While speculative, the stabilizing effect of OT administration on reports of vigor might be related to the (highly understudied, but in the ASD community heavily discussed) phenomenon of ‘autistic burnout’. Individuals with ASD describe ‘autistic burnout’ as an extreme fatigue and inability to meet the demands of everyday life caused by a continuous attempt to mask and/or deal with their ASD symptoms (i.e. sensory disorders, repetitive behaviors (1)). The overall mitigation of repetitive behavior symptoms and increased feelings of social approachability by the OT treatment may therefore have been accompanied with overall higher reports of feeling ‘energetic’, ‘active’, ‘lively’ in the OT group, compared to the PL group. However, considering the exploratory nature of the identified effects, more research is needed to further elucidate the impact of OT treatment on mood states in ASD.

Finally, with respect to the effect of OT on general aspects of quality of life, the current study identified no treatment-specific improvements. To date, evidence on the effects of OT on quality of life is relatively scarce since only two prior studies have addressed this topic. Contrary to our findings, Anagnostou et al. (8) reported an OT-specific improvement in quality of life (socio-emotional section) after a 6-week treatment in adult men with ASD. Watanabe et al. (9) on the other hand, only observed a trend towards improvement in quality of life immediately after a 6-week trial in adult men with ASD. Importantly, recent reviews stated that most individuals with ASD have poor quality of life (note that most studies included children with ASD) (41) or lower quality of life than typically developing adults (42). Note, however, that to date there is no comprehensive ASD-specific quality of life assessment tool validated and consequently, the tools used in the general population (i.e. WHO-QOL) might not be the most adequate to assess quality of life in ASD (and thus changes in quality of life after intervention) (42).

In terms of associations between core autism characteristics and attachment characteristics, our study showed that impairments in social responsiveness and more frequent and/or severe repetitive behaviors were associated with a more avoidant attachment style and with less secure attachment towards significant others (especially the mother). Albeit exploratory, we also showed that, as a group, the individuals with ASD scored higher on avoidant attachment and lower on secure attachment, when compared to a sample of neurotypical individuals. Together, these findings provide indications that - at least to some extent - associations are evident between core autism characteristics and attachment characteristics, a notion that is generally supported by a recent meta-analysis showing an association between the severity of autism characteristics and less secure attachment in children with ASD (22), as well as by other studies showing more insecure attachment towards parents or romantic partners in unmarried (43) or married adults with ASD (44), respectively. However, since research on this topic to date is limited, it currently remains speculative whether the reported feelings of insecure attachment are a result of the social difficulties experienced by individuals with ASD, or conversely, whether difficulties in the social domain are – in part or within a subset of individuals with ASD - a result of a decreased tendency or inability to form secure attachments. Nevertheless, elucidating the interaction between autism symptomology and attachment style may be of particular relevance in the context of OT treatment, since – according to the aforementioned affiliative-motivation hypothesis – especially individuals with a decreased tendency to affiliate (i.e., avoidantly attached individuals) have been proposed to benefit the most from receiving OT treatment (37). The current findings of significant ameliorations in attachment avoidance, but no treatment-specific effects on social responsiveness, are in line with this notion and together suggest that attachment characteristics may be more sensitive for evaluating treatment responses, as compared to evaluations based on core autism characteristics alone. Considering the mixed pattern of effects of OT treatment on core autism symptomatology (e.g. SRS-A, RBS-R, ADOS), it seems of great relevance for future multiple-dose clinical trials with individuals with ASD to additionally include more in-depth characterizations of attachment-related constructs both dimensionally and longitudinally (pre-post treatment). In view of the current observations, these explorations are anticipated to be informative for evaluating and predicting treatment responses, and potentially for delineating patient populations that will benefit the most from a course of OT treatment.

### Limitations

Although the current study provides new insights regarding long-lasting effects of multiple-dose OT treatment in ASD and the relation between autism characteristics and attachment characteristics, several limitations need to be considered. First, although our sample size was comparable to that of prior similar clinical trials, studies with larger samples are warranted. Indeed, considering this is an initial pilot study exploring long-term effects of OT treatment (without correction for multiple comparisons), the findings of long-term improvements in repetitive behaviours; attachment avoidance and vigor mood state should be interpreted with caution. Second, in the current study, participants administered the OT nasal spray once a day (in the morning) while the majority of prior multiple-dose OT studies administered two doses/day (one in the morning and one in the afternoon) (8–10, 12-14, but see 11). While elevated levels of OT have been demonstrated up to 7 hours after a single-dose administration (45), future studies are needed to identify at what point in time the effects of intranasal administration of OT fade out and when OT levels return back to baseline. Also potential interactions with diurnal patterns of endogenous OT levels need to be explored to identify the most optimal dosing and timing of intranasal OT administrations. Third, considering the adopted evaluations were predominantly based on self-report questionnaires, the possibility of subjective bias cannot be ruled out. Participants’ own beliefs about the received treatment, however, were assessed and inclusion of this factor did not modulate the identified treatment effects. Finally, since only adult men with ASD were included, the current observations of beneficial effects of OT treatment cannot be extended to women or children with ASD.

## 5. Conclusions

To conclude, while the four-week, once-daily treatment with OT induced no treatment-specific changes in primary outcome measures of social symptoms in ASD; exploratory analyses of secondary outcomes showed long-lasting, medium to large-sized improvements in repetitive behaviors and a reduction in perceived attachment avoidance, that outlasted the period of actual administration until one month and even one-year post-treatment. Overall, the observation that the OT treatment primarily targeted long-term adaptations in repetitive behaviors and perceived attachment characteristics indicates that these constructs are sensitive for capturing OT treatment effects in adult men with ASD. In line with the central role of the human oxytocinergic system in interpersonal bonding, trust and attachment, the current observations may therefore urge future multiple-dose clinical trials to continue to include characterizations of attachment-related constructs when evaluating the potential of OT treatment for ASD. While the exploratory observations of long-term beneficial effects of OT treatment on repetitive behaviors and perceived attachment avoidance are promising, future studies are warranted to further elucidate the long-term impact of OT treatment.

## Data Availability

Data available upon request.

## 6. List of abbreviations

OT: oxytocin
ASD: autism spectrum disorder
DSM: Diagnostic and Statistical Manual of Mental Disorders
ADOS: Autism Diagnostic Observation Schedule
IU: international units
MRI: magnetic resonance imaging
PL: placebo
POMS: Profile of Mood States
SRS-A: Social Responsiveness Scale
RBS-R: Repetitive Behavior Scale – Revised
SAAM: State Adult Attachment Measure
IPPA: Inventory of Parent and Peer Attachment
WHO-QOL: World Health Organization Quality of Life

## Declarations

### Ethics approval and consent to participate

All participants gave informed written consent in accordance with the ethics approval by the local Ethics Committee for Biomedical Research at the University of Leuven, KU Leuven (s56327).

### Consent for publication

Not applicable.

### Availability of data and materials

The datasets used and/or analysed during the current study are available from the corresponding author on reasonable request.

### Competing interests

The authors declare no competing interests.

### Funding

This research was supported by the Branco Weiss fellowship of the Society in Science - ETH Zurich and by grants from the Flanders Fund for Scientific Research (FWO projects KAN 1506716N, KAN 1521313N, G040112 & G079017N). S.B. is supported by a fund of the Marguerite-Marie Delacroix foundation.

### Authors’ contributions

SB acquired, prepared and analysed the data, and drafted and revised the manuscript. KA and JS designed the study and revised the manuscript. KA also analysed the data. GB and BB provided intellectual contribution to the manuscript and revised the manuscript. All authors read and approved the final manuscript.

## Acknowledgements

We are thankful to all the participating subjects and all the master students who helped with the data collection. We specifically want to acknowledge Claudia Dillen for her help during the start of the clinical trial. We also thank our colleagues of the Leuven Autism Research Consortium.

## Notes

### Competing Interest Statement

The authors have declared no competing interest.

### Clinical Trial

Eudract 2014-000586-45

## References

1. American Psychiatric Association. Diagnostic and Statistical Manual of Mental Disorders (DSM-5®) [Internet]. 5th ed. Arlington VA, editor. American Psychiatric Association; 2013 [cited 2016 Feb 4]. 50–59 p. Available from: https://books.google.com/books?hl=nl&lr=&id=-JivBAAAQBAJ&pgis=1

2. Bakermans-Kranenburg MJ, van IJzendoorn MH. Sniffing around oxytocin: review and meta-analyses of trials in healthy and clinical groups with implications for pharmacotherapy. Transl Psychiatry [Internet]. 2013 Jan 21 [cited 2015 Sep 7];3:e258. Available from: http://dx.doi.org/10.1038/tp.2013.34

3. Bartz JA, Zaki J, Bolger N, Ochsner KN. Social effects of oxytocin in humans: context and person matter. Trends Cogn Sci [Internet]. 2011;15(7):301–9. Available from: http://linkinghub.elsevier.com/retrieve/pii/S1364661311000830

4. Hollander E, Novotny S, Hanratty M, Yaffe R, DeCaria CM, Aronowitz BR, et al. Oxytocin infusion reduces repetitive behaviors in adults with autistic and Asperger’s disorders. Neuropsychopharmacology. 2003;28(1):193–8.

5. Hollander E, Bartz J, Chaplin W, Phillips A, Sumner J, Soorya L, et al. Oxytocin Increases Retention of Social Cognition in Autism. Biol Psychiatry [Internet]. 2007;61(4):498–503. Available from: http://linkinghub.elsevier.com/retrieve/pii/S0006322306007293

6. Guastella AJ, Einfeld SL, Gray KM, Rinehart NJ, Tonge BJ, Lambert TJ, et al. Intranasal oxytocin improves emotion recognition for youth with autism spectrum disorders. Biol Psychiatry [Internet]. 2010 Apr 1 [cited 2015 Dec 16];67(7):692–4. Available from: http://www.sciencedirect.com/science/article/pii/S0006322309011226

7. Andari E, Duhamel J-R, Zalla T, Herbrecht E, Leboyer M, Sirigu A. Promoting social behavior with oxytocin in high-functioning autism spectrum disorders. Proc Natl Acad Sci [Internet]. 2010;107(9):4389–94. Available from: http://www.pnas.org/cgi/doi/10.1073/pnas.0910249107

8. Anagnostou E, Soorya L, Chaplin W, Bartz J, Halpern D, Wasserman S, et al. Intranasal oxytocin versus placebo in the treatment of adults with autism spectrum disorders: a randomized controlled trial. Mol Autism [Internet]. 2012;3(1):16. Available from: http://www.pubmedcentral.nih.gov/articlerender.fcgi?artid=3539865&tool=pmcentrez&rendertype=abstract

9. Watanabe T, Kuroda M, Kuwabara H, Aoki Y, Iwashiro N, Tatsunobu N, et al. Clinical and neural effects of six-week administration of oxytocin on core symptoms of autism. Brain [Internet]. 2015 Nov 1 [cited 2016 Jan 6];138(Pt 11):3400–12. Available from: http://brain.oxfordjournals.org/content/138/11/3400

10. Yamasue H, Okada T, Munesue T, Kuroda M, Fujioka T, Uno Y, et al. Effect of intranasal oxytocin on the core social symptoms of autism spectrum disorder: a randomized clinical trial. Mol Psychiatry [Internet]. 2018;1. Available from: http://www.nature.com/articles/s41380-018-0097-2

11. Dadds MR, MacDonald E, Cauchi A, Williams K, Levy F, Brennan J. Nasal Oxytocin for Social Deficits in Childhood Autism: A Randomized Controlled Trial. J Autism Dev Disord [Internet]. 2014 Mar 26 [cited 2016 Aug 1];44(3):521–31. Available from: http://link.springer.com/10.1007/s10803-013-1899-3

12. Guastella AJ, Gray KM, Rinehart NJ, Alvares G a., Tonge BJ, Hickie IB, et al. The effects of a course of intranasal oxytocin on social behaviors in youth diagnosed with autism spectrum disorders: A randomized controlled trial. J Child Psychol Psychiatry Allied Discip. 2015;4:444–52.

13. Yatawara CJ, Einfeld SL, Hickie IB, Davenport T a, Guastella a J. The effect of oxytocin nasal spray on social interaction deficits observed in young children with autism: a randomized clinical crossover trial. Mol Psychiatry [Internet]. 2016;(August):1–7. Available from: http://www.nature.com.are.uab.cat/mp/journal/vaop/ncurrent/full/mp2015162a.html%5Cnhttp://www.nature.com/doifinder/10.1038/mp.2015.162

14. Parker KJ, Oztan O, Libove RA, Sumiyoshi RD, Jackson LP, Karhson DS, et al. Intranasal oxytocin treatment for social deficits and biomarkers of response in children with autism. Proc Natl Acad Sci [Internet]. 2017;114(30):201705521. Available from: http://www.pnas.org/lookup/doi/10.1073/pnas.1705521114

15. Chevallier C, Kohls G, Troiani V, Brodkin ES, Schultz RT. The social motivation theory of autism. Trends Cogn Sci [Internet]. 2012;16(4):231–8. Available from: http://dx.doi.org/10.1016/j.tics.2012.02.007

16. Shamay-Tsoory SG, Abu-Akel A. The social salience hypothesis of oxytocin. Biol Psychiatry [Internet]. 2015 Aug [cited 2015 Aug 7];79(3):194–202. Available from: http://www.sciencedirect.com/science/article/pii/S0006322315006393

17. Bernaerts S, Prinsen J, Berra E, Bosmans G, Steyaert J, Alaerts K. Long-term oxytocin administration enhances the experience of attachment. Psychoneuroendocrinology. 2017;78:1–9.

18. De Dreu CKW. Oxytocin modulates the link between adult attachment and cooperation through reduced betrayal aversion. Psychoneuroendocrinology [Internet]. 2012 Jul [cited 2015 Dec 9];37(7):871–80. Available from: http://www.sciencedirect.com/science/article/pii/S0306453011003064

19. Bartz J a., Lydon JE, Kolevzon a., Zaki J, Hollander E, Ludwig N, et al. Differential Effects of Oxytocin on Agency and Communion for Anxiously and Avoidantly Attached Individuals. Psychol Sci [Internet]. 2015;1–10. Available from: http://pss.sagepub.com/lookup/doi/10.1177/0956797615580279

20. Buchheim A, Heinrichs M, George C, Pokorny D, Koops E, Henningsen P, et al. Oxytocin enhances the experience of attachment security. Psychoneuroendocrinology [Internet]. 2009;34(9):1417–22. Available from: http://www.pubmedcentral.nih.gov/articlerender.fcgi?artid=3138620&tool=pmcentrez&rendertype=abstract

21. Rutgers AH, Bakermans-Kranenburg MJ, Van Ijzendoorn MH, Van Berckelaer-Onnes IA. Autismand attachment: ameta-analytic review. J Child Psychol Psychiatry. 2004;45(6):1123–34.

22. Teague SJ, Gray KM, Tonge BJ, Newman LK. Attachment in children with autism spectrum disorder: A systematic review. Res Autism Spectr Disord [Internet]. 2017;35:35–50. Available from: http://dx.doi.org/10.1016/j.rasd.2016.12.002

23. Lord C, Rutter M, Goode S, Heemsbergen J, Jordan H, Mawhood L, et al. Autism diagnostic observation schedule: A standardized observation of communicative and social behavior. J Autism Dev Disord [Internet]. 1989 Jun [cited 2018 Jun 11];19(2):185–212. Available from: http://link.springer.com/10.1007/BF02211841

24. Lord C, DiLavore P, Gotham K, Guthrie W, Luyster R, Risi S, et al. Autism diagnostic observation schedule: ADOS-2 [Internet]. 2nd ed. Los Angeles: Western Psychological Services; 2012 [cited 2019 Jun 19]. Available from: https://www.worldcat.org/title/autism-diagnostic-observation-schedule-ados-2/oclc/851410387

25. Wechsler D, Coalson D, Raiford S. WAIS IV Technical and Interpretive Manual. San Antonio: NCS Pearson; 2008.

26. Domes G, Heinrichs M, Michel A, Berger C, Herpertz SC. Oxytocin improves “mind-reading” in humans. Biol Psychiatry [Internet]. 2007 Mar 15 [cited 2015 Jun 1];61(6):731–3. Available from: http://www.sciencedirect.com/science/article/pii/S0006322306009395

27. Guastella AJ, Mitchell PB, Dadds MR. Oxytocin increases gaze to the eye region of human faces. Biol Psychiatry [Internet]. 2008 Jan 1 [cited 2016 Jan 13];63(1):3–5. Available from: http://www.sciencedirect.com/science/article/pii/S0006322307006178

28. Gordon I, Vander Wyk BC, Bennett RH, Cordeaux C, Lucas M V., Eilbott JA, et al. Oxytocin enhances brain function in children with autism. Proc Natl Acad Sci [Internet]. 2013;110(52):20953–8. Available from: http://www.pnas.org/cgi/doi/10.1073/pnas.1312857110

29. Guastella AJ, MacLeod C. A critical review of the influence of oxytocin nasal spray on social cognition in humans: Evidence and future directions. Horm Behav [Internet]. 2012;61(3):410–8. Available from: http://linkinghub.elsevier.com/retrieve/pii/S0018506X12000037

30. Bernaerts S, Berra E, Wenderoth N, Alaerts K. Influence of oxytocin on emotion recognition from body language: A randomized placebo-controlled trial. Psychoneuroendocrinology. 2016;72.

31. Constantino JN. Social Responsiveness Scale-Adult Research Version. Lon Angeles, CA: Western Psychological Services; 2005.

32. Lam KSL, Aman MG. The repetitive behavior scale-revised: Independent validation in individuals with autism spectrum disorders. J Autism Dev Disord. 2007;37(5):855–66.

33. Gillath O, Hart J, Noftle EE, Stockdale GD. Development and validation of a state adult attachment measure (SAAM). J Res Pers [Internet]. 2009 Jun [cited 2016 Feb 15];43(3):362–73. Available from: http://www.sciencedirect.com/science/article/pii/S0092656608001694

34. Armsden GC, Greenberg MT. The inventory of parent and peer attachment: Individual differences and their relationship to psychological well-being in adolescence. J Youth Adolesc [Internet]. 1987;16(5):427–54. Available from: http://dx.doi.org/10.1007/BF02202939

35. The Whoqol Group. The World Health Organization quality of life assessment (WHOQOL): Development and general psychometric properties. Soc Sci Med [Internet]. 1998 Jun 15 [cited 2018 Jun 13];46(12):1569–85. Available from: https://www.sciencedirect.com/science/article/pii/S0277953698000094?via%3Dihub

36. Cohen J. Statistical Power Analysis for the Behavioral Sciences. 2nd ed. New Jersey: Lawrence Erlbaum Associates; 1988.

37. Bartz JA. Oxytocin and the Pharmacological Dissection of Affiliation. Curr Dir Psychol Sci [Internet]. 2016;25(2):104–10. Available from: http://cdp.sagepub.com/lookup/doi/10.1177/0963721415626678

38. Rodgers J, Glod M, Connolly B, McConachie H. The Relationship Between Anxiety and Repetitive Behaviours in Autism Spectrum Disorder. J Autism Dev Disord [Internet]. 2012 Nov 17 [cited 2018 Jul 25];42(11):2404–9. Available from:http://link.springer.com/10.1007/s10803-012-1531-y

39. Lang M, Krátký J, Shaver JH, Jerotijević D, Xygalatas D. Effects of Anxiety on Spontaneous Ritualized Behavior. Curr Biol [Internet]. 2015 Jul 20 [cited 2018 Jul 25];25(14):1892–7. Available from: https://www.sciencedirect.com/science/article/pii/S0960982215006521?via%3Dihub

40. Masi A, Lampit A, Glozier N, Hickie IB, Guastella AJ. Predictors of placebo response in pharmacological and dietary supplement treatment trials in pediatric autism spectru disorder: A meta-analysis. Transl Psychiatry [Internet]. 2015;5(9):e640-9. Available from: http://dx.doi.org/10.1038/tp.2015.143

41. Chiang HM, Wineman I. Factors associated with quality of life in individuals with autism spectrum disorders: A review of literature. Res Autism Spectr Disord [Internet]. 2014;8(8):974–86. Available from: http://dx.doi.org/10.1016/j.rasd.2014.05.003

42. Ayres M, Parr JR, Rodgers J, Mason D, Avery L, Flynn D. A systematic review of quality of life of adults on the autism spectrum. Autism. 2017;

43. Taylor EL, Target M, Charman T. Attachment in adults with high-functioning autism. Attach Hum Dev. 2008;10(2):143–63.

44. Lau W, Peterson CC. Adults and children with Asperger syndrome: Exploring adult attachment style, marital satisfaction and satisfaction with parenthood. Res Autism Spectr Disord [Internet]. 2011 Jan 1 [cited 2018 Jun 20];5(1):392–9. Available from: https://www.sciencedirect.com/science/article/pii/S1750946710000887

45. van Ijzendoorn MH, Bhandari R, van der Veen R, Grewen KM, Bakermans-Kranenburg MJ. Elevated Salivary Levels of Oxytocin Persist More than 7 h after Intranasal Administration. Front Neurosci [Internet]. 2012 [cited 2019 Mar 20];6:174. Available from: http://www.ncbi.nlm.nih.gov/pubmed/23233832

